# DNA methylation patterns define subtypes of differentiated follicular cell-derived thyroid neoplasms: an unsupervised machine learning approach

**DOI:** 10.1101/2022.12.19.22283657

**Authors:** Vicente Rodrigues Marczyk, Mariana Recamonde-Mendoza, Ana Luiza Maia, Iuri Martin Goemann

## Abstract

Alterations in DNA methylation patterns are a frequent finding in cancer. Methylation aberrations can drive tumorigenic pathways and serve as potential biomarkers. The role of epigenetic alterations in thyroid cancer is still poorly understood Here, we analyzed methylome data of a total of 810 thyroid samples (n=256 for discovery and n=554 for validation), including benign and malignant follicular cell-derived thyroid neoplasms, as well as normal thyroid tissue. In the discovery phase, we employed an unsupervised machine-learning method to search for methylation patterns. We found evidence supporting the existence of three distinct methylation subtypes: a normal-like, a hypermethylated follicular-like, and a hypomethylated papillary-like cluster. Follicular adenomas, follicular carcinomas, oncocytic adenomas, oncocytic carcinomas, and NIFTP samples were grouped within the follicular-like cluster, indicating that these pathologies shared numerous epigenetic alterations, with a predominance of hypermethylation events. Conversely, classic papillary thyroid carcinomas (PTC) and tall cell PTC formed a separate subtype characterized by the predominance of hypomethylated positions. Interestingly, follicular variant papillary thyroid carcinomas (FVPTC) were as likely to be classified as follicular-like or PTC-like during the discovery phase, indicating a heterogeneous group likely to be formed by at least two distinct diseases. In the validation phase, we found that FVPTC with follicular-like methylation patterns were enriched for RAS mutations. In contrast, FVPTC with PTC-like methylation patterns were enriched for BRAF and RET alterations. Our data provide novel insights into the epigenetic alterations of thyroid tumors. Since the classification method relies on a fully unsupervised machine learning approach for subtype discovery, our results offer a robust background to support the classification of thyroid neoplasms based on methylation patterns.

## Introduction

Follicular cell-derived thyroid neoplasms are the most frequent endocrine malignancy worldwide^1^. These tumors can be classified into benign lesions, low-risk neoplasms, and malignant neoplasms^2^. However, diagnosis can be challenging, and many lesions can only be diagnosed after surgical excision. A better understanding of tumor genetics has given new perspectives on classifying thyroid neoplasms. However, studies have focused on genetic rather than epigenetic alterations such as methylation, as recently reviewed by Zafon *et al*^3^. Most studies evaluating the thyroid cancer DNA methylome used array-based technologies. Nevertheless, their conclusions were limited either by the restricted number of samples or by the underrepresentation of many histological subtypes. To date, no study has employed a robust framework to classify thyroid neoplasms based on their methylation patterns. Our study aims to analyze methylation patterns among various follicular cell-derived thyroid neoplasms and identify the existence of disease clusters that can be robust and reliably identified using methylation microarray data through unsupervised consensus clustering.

Unsupervised clustering is a powerful method to identify disease subtypes, and it has been extensively employed in cancer research^4,5^. However, clustering techniques frequently encounter evidence for clustering even in data where no real clusters exist, leading to equivocal interpretation about the existence of subtypes within a homogenous group^6^. Additionally, all commonly used clustering methods, such as K-means (KM), partitioning around medoids (PAM), or even hierarchical clustering (HC), require the researcher to at some point specify the number of clusters into which to split the data. This decision is complex since the number of categories is usually unknown beforehand. Monte Carlo reference-based consensus clustering (M3C) is a recently developed approach that robustly addresses the issue of identifying the true K number of clusters^7^. M3C is a consensus clustering approach (i.e., it resamples the data multiple times to evaluate how often a pair of samples clusters together) built upon the original Monti algorithm^8^ using a robust metric, the proportion of ambiguous clustering (PAC)^6^, and a reference distribution^7^.

Here, we demonstrate that consensus clustering can identify distinct methylation patterns and help us understand thyroid tumor biology. We propose a novel way to classify follicular cell-derived thyroid neoplasms based on their methylome.

## Methods

### Search strategy and eligibility of studies

We performed a systematic search until September 2022 using the Gene Expression Omnibus (GEO) for datasets with available DNA methylation data on thyroid cancer and benign/normal thyroid tissue analyzed by array methods using the following query (“thyroid neoplasms”[MeSH Terms] OR thyroid cancer[All Fields]) AND “Homo sapiens”[porgn] AND (“Methylation profiling by array”[Filter] OR “Methylation profiling by genome tiling array”[Filter] OR “Methylation profiling by SNP array”[Filter]). The description for each resulting dataset was read in detail to evaluate whether it was suitable for inclusion. Datasets originating from *in vitro* studies or generated from blood samples were omitted. Rare histological subtypes (< 5 samples in total), anaplastic thyroid carcinomas, and medullary thyroid carcinomas were not included. The nomenclature of thyroid neoplasms was maintained as originally described in each study.

### Data analyses

All analyses were conducted in R version 4.1.3. Raw data were retrieved from GEO in the form of *idat* files using the *GEOquery* package^9^. Methylation data preprocessing was performed using the *minfi* package (version 1.40.0)^10^. Data normalization was performed using single-sample Noob normalization, which is advantageous when working across array types with large datasets arriving in batches^11^. Probes were filtered out to exclude those of low quality, probes mapped to the X and Y chromosomes, probes with SNPs, and cross-reactive probes. We inspected our data for the presence of batch effects through dimensionality reduction techniques, finding only minor batch effects present within our data. Since the datasets included in our analyses were highly unbalanced regarding tumor subtypes, batch correction retaining group differences may introduce a strong bias, deflating p values and leading to false discoveries^12^. Therefore, we chose a more conservative approach, using our data without batch correction for all downstream analyses. For completeness, we also ran all our analyses adjusting for the study batch using the empirical Bayes method from ComBat^13^ and retaining the differences between tumor subtypes. We found no important changes in the results, reinforcing that batch effects were not responsible for most of the variation within our data.

### Unsupervised machine learning

We employed a Monte Carlo reference-based consensus clustering (M3C)^7^ approach using the *M3C* package (version 1.16.0) with default parameters unless otherwise specified. We conducted 100 Monte Carlo iterations and 100 inner replications. The optimal number of clusters (K) was defined based on two metrics: the Relative Cluster Stability Index (RCSI) and the Monte Carlo p value. Consensus clustering was performed using beta-values with the largest variance, ranging from 5,000 to 20,000 features included. We executed the consensus clustering algorithm using spectral clustering, which can deal with anisotropic clusters, unequal variances, and non-Gaussian shapes^7^. We employed the proportion of ambiguous clustering (PAC) as the objective function for M3C clustering. M3C was also used for t-SNE plots using default parameters. Hierarchical clustering with the 5,000 most variable beta-values was performed for visualization using the *dendextend* package (version 1.16.0) with Euclidean distances and Ward’s linkage.

### Validation in an independent dataset

Data from The Cancer Genome Project (TCGA)^14^ were retrieved as *idat* files from the GDC Data Portal. Data preprocessing using *minfi* was performed in the same manner as for GEO datasets. Samples from metastatic sites or rare histological subtypes (< 5 samples in total) were not included. We employed the nearest shrunken centroids method^15^ available in the *pamr* package (version 1.56.1) to train a classifier using the 5,000 most variable beta-values in our discovery dataset. The classifier was trained based on cluster assignment by M3C spectral clustering with the same 5,000 features and worked by assigning new samples to the class with the minimum distance between the sample and the shrunken centroid obtained in the clustering analysis. Our classifier was unaware of the sample histological subtype. We tested our classifier using TCGA methylation data to inspect the cluster assignment for each histological subtype. Mutational data were retrieved from cBioPortal^16^, and oncoprint visualizations were constructed using the *ComplexHeatmap* package (version 2.10.0).

### Differential methylation analysis

We performed differential methylation analysis (DMA) using the *DMRCate* package (version 2.8.5)^17^. We conducted DMA to identify differentially methylated CpG loci and differentially methylated regions among clusters. We adjusted p values for multiple comparisons using the BenjaminiãHochberg method. We set our threshold for differential methylation at a minimum beta-value difference of 0.2 and an adjusted p value < 0.05.

### Gene set and pathway analysis

We conducted gene set analysis (GSA) to better understand the biological significance of differentially methylated CpG loci employing Reactome^18^ gene sets for signal transduction pathways (R-HSA-162582) and diseases of signal transduction (R-HSA-5663202) with 60 to 300 genes. We conducted GSA using robust rank aggregation and overrepresentation analysis using the *methylGSA* package^19^.

## Results

### Thyroid cancer array-based methylation studies

We identified a total of 13 studies, of which four were included (Supplementary Figure 1): GSE121377^20^, GSE77804^21^, GSE97466^22^, and GSE197860^23^. An additional study from GEO (GSE53051^24^) was identified from references and was included in our analysis. All five datasets employed Illumina platforms, with two studies using the EPIC array and three employing the 450K array. We included a total of 256 thyroid samples as follows: 70 classic papillary thyroid carcinomas (cPTC), 22 follicular thyroid carcinomas (FTC), 21 follicular variant papillary thyroid carcinomas (FVPTC), 16 oncocytic carcinomas (OC), 16 oncocytic adenomas (OA), 16 follicular adenomas (FA), and 6 noninvasive follicular thyroid neoplasms with papillary-like nuclear features (NIFTP) (Supplementary Table 1).

### Determining the true number of clusters using Monte Carlo consensus clustering

After data preprocessing, we obtained a matrix of normalized methylation beta-values for 256 samples and 332,290 genomic positions for subsequent analyses. Ideally, the optimal number of clusters should be the number K that maximizes the relative cluster stability index (RCSI) and the -log_2_ Monte Carlo p value. Our results indicated the existence of three (K=3) methylation clusters within our dataset, irrespective of whether we employed 5, 10, or 20 thousand features for clustering (Figure 1 and Supplementary Figure 2). To evaluate whether our hypothesis of K=3 methylation clusters was reasonable, we performed a visual inspection to assess cluster separation using t-SNE (Figure 2), which supported K=3 as the optimal number of clusters. Next, we assessed cluster assignment consistency by comparing the cluster assignment for a given sample as we varied the number of features employed. Regardless of whether we used 5, 10, or 20 thousand features for clustering, 243 out of 256 samples (94.9%) were always assigned to the same subtype, demonstrating consistent results.

**Figure 1:**
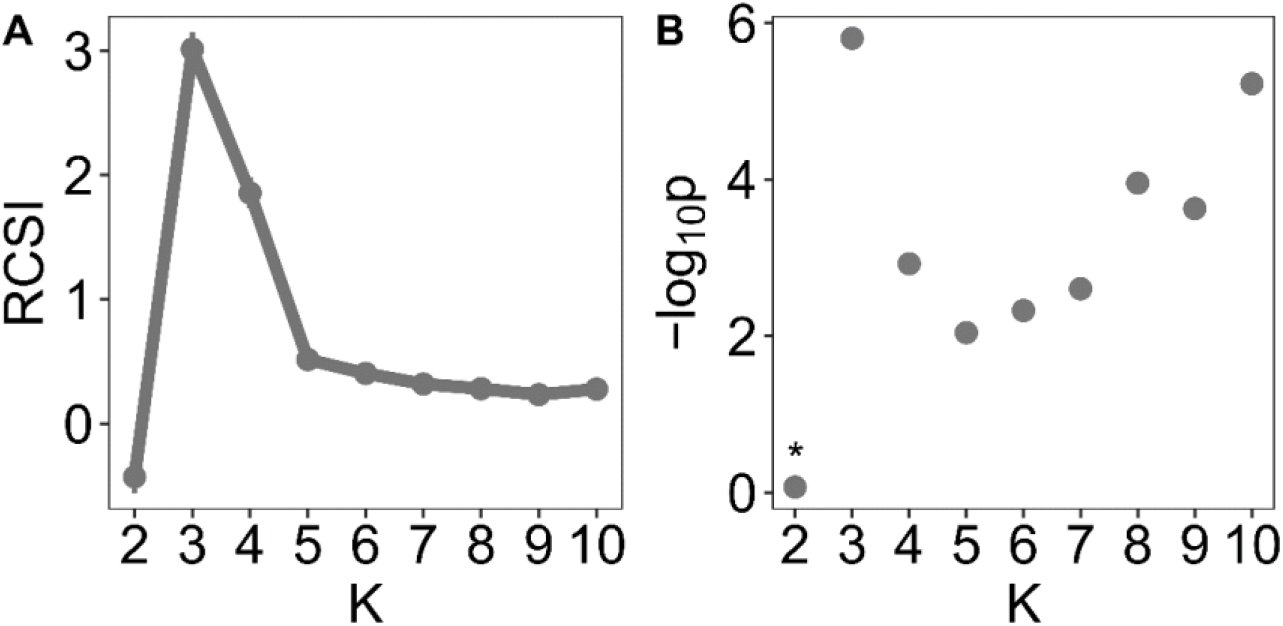
Stability metrics (RCSI and Monte Carlo p value) obtained from spectral consensus clustering using the 5,000 most variable beta-values. **Abbreviations:** RCSI, Relative Cluster Stability Index. *Not statistically significant.

**Figure 2:**
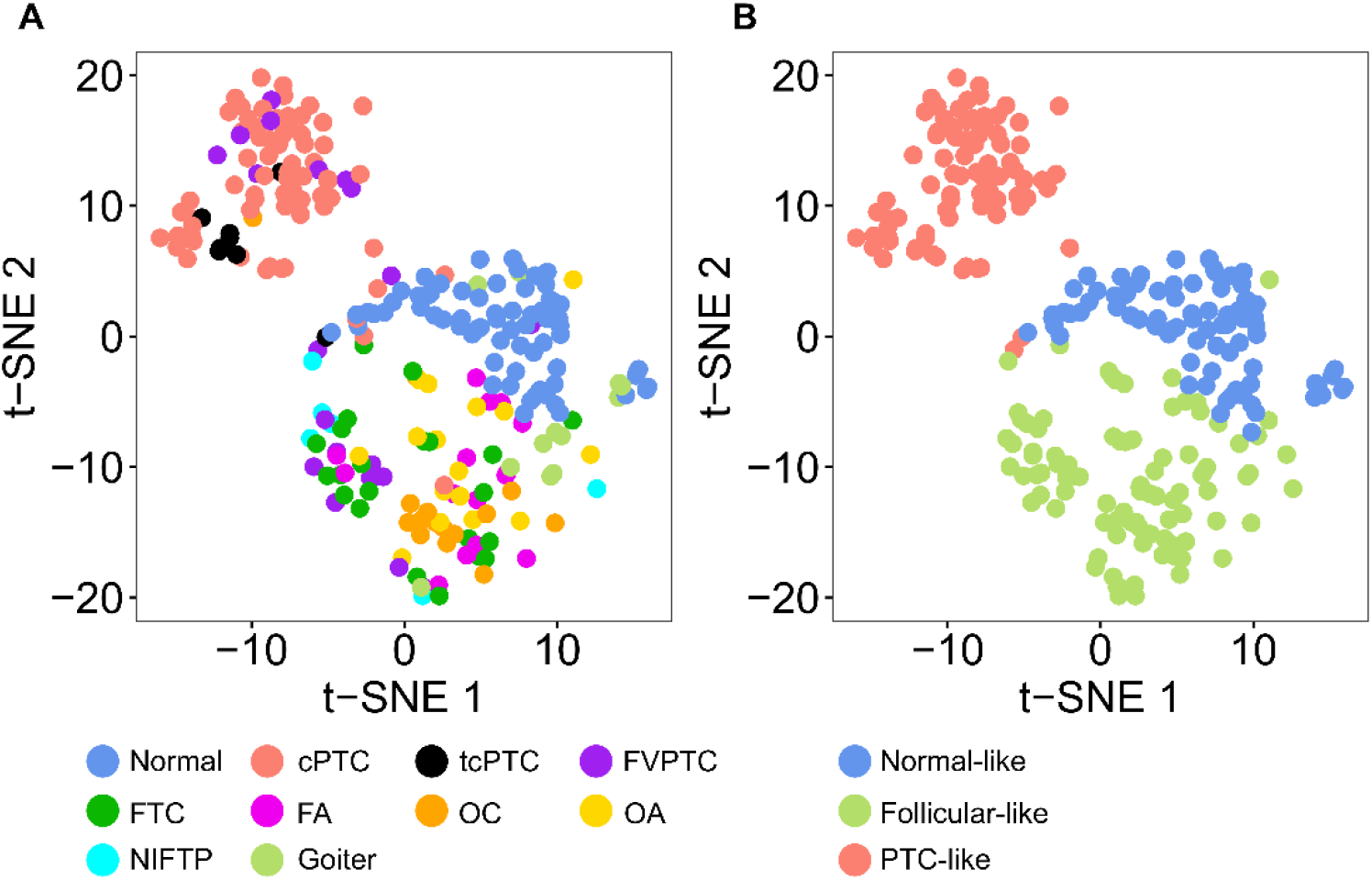
t-SNE plots were employed to inspect cluster separation. Samples were colored (A) according to histological subtype or (B) according to cluster assignment based on spectral clustering with 5,000 features. **Abbreviations:** cPTC, classic papillary thyroid carcinoma; FA, follicular adenoma; FVPTC, follicular variant papillary thyroid carcinoma; FTC, follicular thyroid carcinoma; OA, oncocytic adenoma; OC, oncocytic carcinoma; NIFTP: noninvasive follicular thyroid neoplasm with papillary-like nuclear features; tcPTC, tall cell papillary thyroid carcinoma; t-SNE, t-distributed stochastic neighbor embedding.

### Association between methylation clusters and histological subtypes

From the t-SNE plots, we observed an association between cluster assignment and histological subtypes (Figure 2). For visualization purposes, we performed conventional hierarchical clustering into K=3 clusters (Figure 3). Cluster 1 (“normal-like”) was composed primarily of normal thyroid tissue and goiter samples; cluster 2 (“follicular-like”) was composed of FA, FTC, OA, OC, and NIFTP; and cluster 3 (“PTC-like”) was composed predominantly of cPTC and tcPTC (Figure 3).

**Figure 3:**
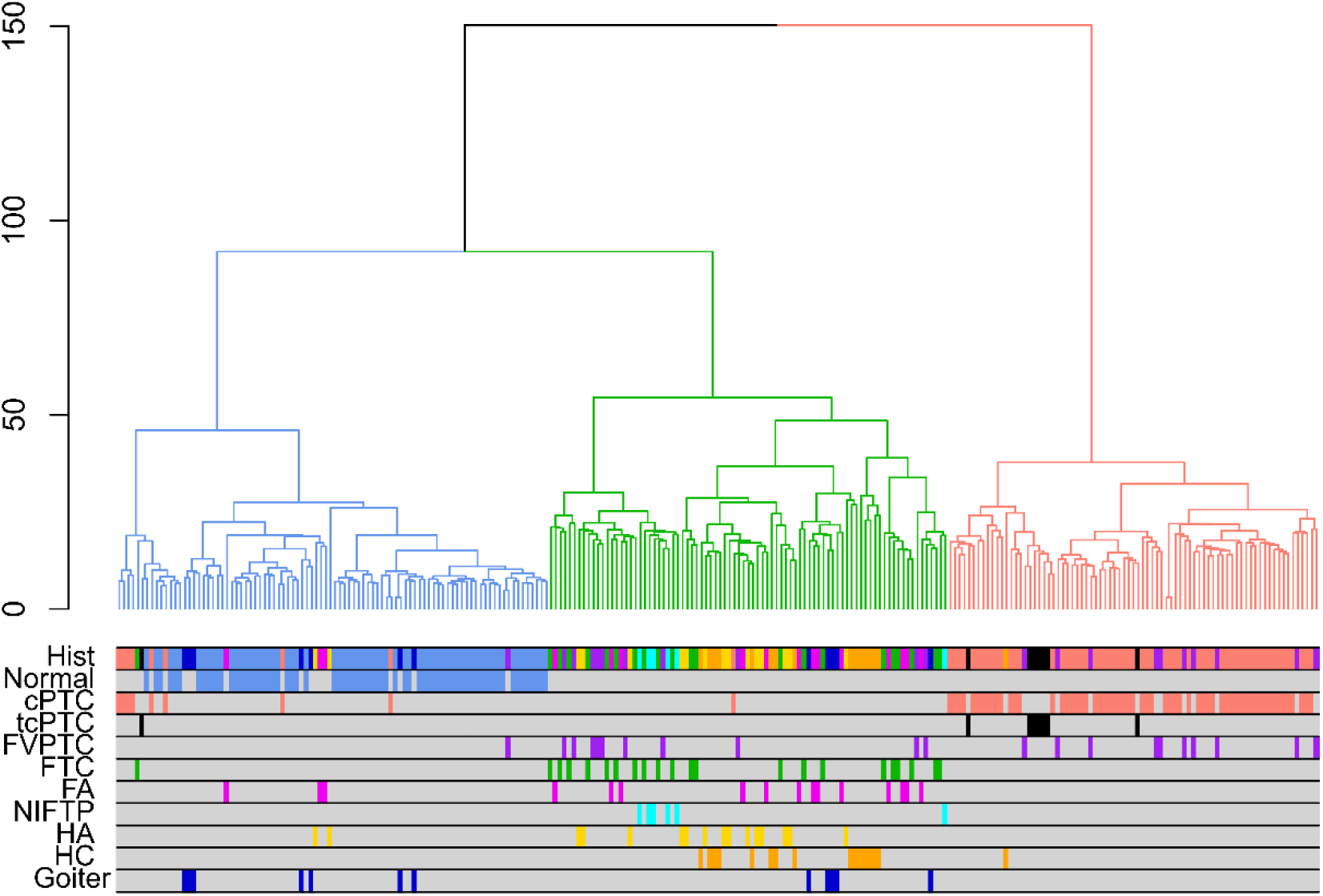
Hierarchical clustering employing the 5,000 most variable beta-values was performed to cluster samples into K=3 groups. Euclidean distances and Ward linkage were employed. **Abbreviations:** cPTC, classic papillary thyroid carcinoma; FA, follicular adenoma; FVPTC, follicular variant papillary thyroid carcinoma; FTC, follicular thyroid carcinoma; OA, oncocytic adenoma; OC, oncocytic carcinoma; NIFTP: noninvasive follicular thyroid neoplasm with papillary-like nuclear features; tcPTC, tall cell papillary thyroid carcinoma.

Consensus clustering results also demonstrated a significant association between methylation patterns and histological subtypes (Fisher’s exact p < 0.001) (Table 2). Classic PTC and tcPTC were preferentially assigned to the PTC-like cluster, with 91.4% cPTC and 100% tcPTC assigned to this group. There was also a remarkable aggregation of samples for neoplasms exhibiting follicular or oncocytic patterns. All FA and all OA were assigned to the follicular-like cluster. For their malignant counterparts, 100% FTC and 93.8% OC were also placed within this subtype. It is noteworthy that, although represented in a small number, 6 out of 6 (100%) NIFTP samples were placed within the follicular-like cluster rather than in the PTC-like cluster. Unlike all other histologic subtypes previously discussed, for which preferential clustering was evident, FVPTC did not behave as a homogeneous group, and samples were as likely to be classified as follicular-like (47.6%) as they were to be classified as PTC-like (42.9%) (Table 2). Accordingly, FVPTC samples did not cluster together on the t-SNE plot (Figure 2) or on the hierarchical clustering dendrogram (Figure 3), suggesting that this histological subtype might, in fact, represent two distinct diseases grouped as one.

**Table 2:** Cluster assignment for each histological subtype Cluster assignment for each histological subtype based on spectral consensus clustering with the 5,000 most variable beta-values. Row-wise p values were calculated using a multinomial exact test of goodness-of-fit with the null hypothesis that each tumor subtype was equally likely to be assigned to any of the clusters. **Abbreviations:** cPTC, classic papillary thyroid carcinoma; FA, follicular adenoma; FVPTC, follicular variant papillary thyroid carcinoma; FTC, follicular thyroid carcinoma; OA, oncocytic adenoma; OC, oncocytic carcinoma; NIFTP: noninvasive follicular thyroid neoplasm with papillary-like nuclear features; tcPTC, tall cell papillary thyroid carcinoma. *Not statistically significant.

|  |  | Normal-like | Follicular-like | PTC-like | p-value |
| --- | --- | --- | --- | --- | --- |
| Normal | n | 69 | 0 | 0 | < 0.001 |
|  | % | (100) | (0) | (0) |  |
| cPTC | n | 5 | 1 | 64 | < 0.001 |
|  | % | (7.1) | (1.4) | (91.4) |  |
| tcPTC | n | 0 | 0 | 8 | < 0.001 |
|  | % | (0) | (0) | (100) |  |
| FVPTC | n | 2 | 10 | 9 | 0.052* |
|  | % | (9.5) | (47.6) | (42.9) |  |
| FTC | n | 0 | 22 | 0 | < 0.001 |
|  | % | (0) | (100) | (0) |  |
| FA | n | 0 | 16 | 0 | < 0.001 |
|  | % | (0) | (100) | (0) |  |
| OC | n | 0 | 15 | 1 | < 0.001 |
|  | % | (0) | (93.8) | (6.2) |  |
| OA | n | 0 | 16 | 0 | < 0.001 |
|  | % | (0) | (100) | (0) |  |
| NIFTP | n | 0 | 6 | 0 | 0.004 |
|  | % | (0) | (100) | (0) |  |
| Goiter | n | 6 | 6 | 0 | 0.031 |
|  | % | (50.0) | (50.0) | (0) |  |

### Validation using an independent dataset

We validated our findings using TCGA samples as an independent external dataset. Samples were assigned a cluster exclusively based on their methylation pattern compared to the three previously defined clusters, without any information about histological subtypes. The results were in accordance with the discovery phase (Table 3 and Supplementary Figure 3), suggesting that true methylation clusters were correctly identified. Our model classified 94.6% of normal thyroid tissue within the normal-like cluster. All 38 (100%) tcPTC and 83.9% cPTC were assigned to the PTC-like cluster. Interestingly, FVPTC were predominantly located within the follicular-like cluster (72.4%) and less frequently in the PTC-like cluster (24.8%). For TCGA samples, we also had mutational data available, so we tested whether FVPTC assigned to the follicular-like group differed from FVPTC assigned to the PTC-like group in terms of DNA mutations (Figure 4). We observed that follicular-like FVPTC were characterized by a higher frequency of RAS mutations (43.4% *vs*. 11.5%, Fisher exact p < 0.001), whereas PTC-like FVPTC were associated with alterations of BRAF (53.8% *vs*. 5.3%, Fisher exact p = 0.004) and RET (15.4% *vs*. 0%, Fisher exact p = 0.004). Other mutations (TG, EIF1AX) and fusions (THADA, PAX8, NTRK3) frequently associated with thyroid carcinomas were also inspected, but their low frequency did not provide sufficient power for formal testing (Figure 4).

**Table 3:** Cluster assignment for TCGA samples Cluster assignment for each histological subtype in the TCGA dataset. Cluster assignment was performed using a classifier based on the nearest shrunken centroids calculated from the discovery dataset. Row-wise p values were calculated using a multinomial exact test of goodness-of-fit with the null hypothesis that each tumor subtype was equally likely to be assigned to any of the clusters. **Abbreviations:** cPTC, classic papillary thyroid carcinoma; FVPTC, follicular variant papillary thyroid carcinoma; tcPTC, tall cell papillary thyroid carcinoma; TCGA, The Cancer Genome Atlas.

|  | Normal-like |  | Follicular-like |  | PTC-like |  | p-value |
| --- | --- | --- | --- | --- | --- | --- | --- |
|  | n | (%) | n | (%) | n | (%) |  |
| Normal | 53 | (94.6) | 3 | (5.4) | 0 | 0 | <0.001 |
| cPTC | 13 | (3.7) | 44 | (12.4) | 298 | (83.9) | <0.001 |
| tcPTC | 0 | 0 | 0 | 0 | 38 | -100 | <0.001 |
| FVPTC | 3 | (2.9) | 76 | (72.4) | 26 | (24.8) | <0.001 |

**Figure 4:**
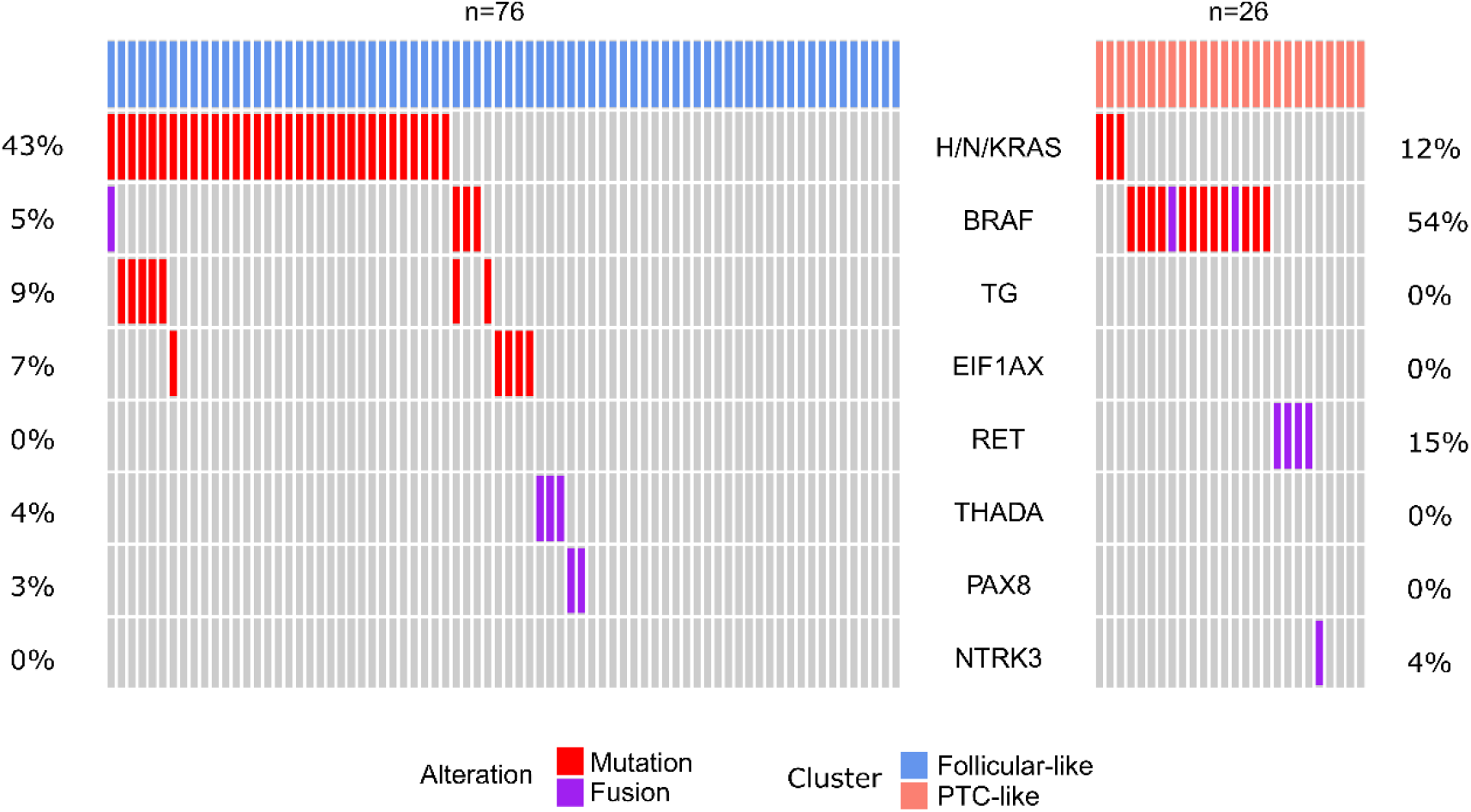
Oncoprint representation comparing the mutational profile of FVPTC with a follicular-like methylation pattern (left) and FVPTC with a PTC-like methylation pattern (right). Each individual column represents a single sample, and rows represent genes. **Abbreviations:** PTC, papillary thyroid carcinoma; FVPTC, follicular variant papillary thyroid carcinoma.

### Differential methylation analysis

Next, we performed differential methylation analysis to better understand the biological differences between these clusters identified during the discovery phase. Using the normal-like cluster as our reference group, we observed that follicular-like samples exhibited 1,757 differentially methylated positions, of which 89.1% were hypermethylated (Table 4). Conversely, PTC-like samples exhibited 3,241 differentially methylated sites, with a predominance of hypomethylated positions (87.5%) (Table 4). It is known, however, that most biologically significant differences in methylation affect genomic regions rather than isolated CpG loci. Therefore, we also conducted a differential methylation analysis at the genomic region level. Once again, we observed that follicular-like samples were marked mainly by hypermethylation events, whereas PTC-like samples exhibited more hypomethylated regions (Table 4). A detailed description of these genomic regions along with a complete list of differentially methylated positions is available in the Supplementary Material. We also investigated whether the differentially methylated positions mapped to genes associated with specific signaling pathways. For PTC-like samples, we found a highly significant association with the VEGF, PI3K/AKT, and MAPK signaling pathways (adjusted p value <0.001 for all three pathways) (Supplementary Table 2). For follicular-like samples, differentially methylated sites were associated with a greater variety of pathways, including VEGF, MAPK, NOTCH, and TGFB (adjusted p value <0.001) (Supplementary Table 3).

**Table 4:** Differentially methylated genomic positions and regions in comparison to the normal-like cluster

|  |  | Differentially methylated positions |  |  | Differentially methylated regions |  |
| --- | --- | --- | --- | --- | --- | --- |
|  |  | Follicular-like | PTC-like |  | Follicular-like | PTC-like |
| **Hypermethylated** | n | 1565 | 405 | n | 55 | 9 |
|  | (%) | (89.1) | (12.5) | (%) | (96.5) | (6.6) |
| **Hypomethylated** | n | 192 | 2836 | n | 2 | 127 |
|  | (%) | (10.9) | (87.5) | (%) | (3.5) | (93.4) |
| **Total** | n | 1757 | 3241 | n | 57 | 136 |

## Discussion

Our study offers the most comprehensive evaluation of the thyroid neoplasm methylome to date. Using an unsupervised machine-learning method, we identified the existence of three thyroid methylation subtypes: normal-like, hypermethylated follicular-like, and hypomethylated PTC-like.

Previous studies have shown methylome dysregulation in thyroid neoplasms, mostly using array-based methods^3^. Studies focused on PTC have consistently reported that these tumors are marked by hypomethylation events^14,22,25,26^, which is consistent with our results. FTC has been far less studied, with some studies reporting hypermethylation in this disease^22,27,28^, which is in accordance with our findings. To date, the most comprehensive study has been reported by Bisaro dos Reis *et al*.^22^, who analyzed 141 thyroid samples of various types, including normal, benign, and cancer samples. The authors proposed the existence of methylation clusters based on unsupervised hierarchical clustering, a classification that partially overlaps ours. Their study, however, had a very limited number of histological subtypes, such as FTC (n=8), OC (n=2), OA (n=3), and TCPTC (n=1). Additionally, their clustering strategy did not formally address the issue of finding the true number of clusters. Additionally, in the TCGA study,^14^ a methylome assessment of a large cohort of patients with thyroid cancer was also performed. Nevertheless, the data were limited to PTC and normal samples, offering a deep understanding of this histological subtype but precluding more general conclusions about follicular cell-derived thyroid neoplasms as a broader group.

Here, we show that cPTC and tcPTC have a distinct methylation pattern marked by hypomethylation events, defining the PTC-like methylation subtype. Conversely, samples exhibiting follicular or oncocytic histology have a pattern marked by hypermethylated sites, defining the follicular-like methylation subtype. It is noteworthy that follicular and oncocytic neoplasms were grouped irrespective of whether they were benign (follicular and oncocytic adenomas) or malignant tumors (follicular and oncocytic carcinomas), supporting the hypothesis that follicular adenomas and follicular carcinomas are likely to be entities within the same disease continuum. A common origin for both diseases is supported by DNA mutation analysis showing that both are characterized by RAS mutation, with follicular adenomas exhibiting these mutations in 20-40% of the cases, whereas follicular carcinomas present RAS mutations at a higher frequency (40-50%)^29–31^. Follicular adenomas and carcinomas also share multiple transcriptional similarities at the RNA and miRNA levels, favoring a common origin for both pathologies^32^.

Interestingly, our results indicate that FVPTC does not exhibit a homogenous methylation pattern, suggesting that this histological subtype might be a heterogeneous group formed by more than one disease rather than a tumor subtype itself. Most FVPTC have a follicular-like methylation pattern, suggesting that these tumors are more closely related to FTC than to PTC. These follicular-like FVPTC also show a predominance of RAS mutations, reinforcing that they are closer to FTC. Conversely, a smaller fraction of FVPTC has a PTC-like methylome, suggesting that they are truly related to PTC. These PTC-like FVPTC were enriched for BRAF and RET alterations, strengthening this hypothesis. Our results are in accordance with previous reports describing FVPTC as a heterogeneous entity. Encapsulated FVPTC is marked by RAS mutations, resembling FTC, whereas infiltrative tumors are characterized by BRAF mutations, resembling PTC^2,33^. The TCGA authors also discuss the classification of FVPTC and suggest that these tumors be classified alongside FTC, since they are marked by RAS mutations and arm-level genetic alterations^14^. Our data offer new insights from an epigenetic perspective to help understand and classify FVPTC more precisely.

Our study has some limitations that must be recognized. Our results were derived from samples collected from different cohorts of patients, processed differently, and assessed using two distinct methylation arrays (450K and EPIC). This concern is diminished given that we could reproduce our results in a large independent cohort of patients not used during the discovery phase. Our study does not provide information about less frequent histological subtypes since they were not sufficiently represented within our dataset. Additionally, we are dependent on the reported original histological classification of samples, which might be inaccurate, especially in light of the new WHO classification of thyroid neoplasms.

In conclusion, our data provide broad insights into the major epigenetic alterations present within thyroid tumors. Using a fully unsupervised machine learning method for subtype discovery, our results offer a robust background to support the classification of thyroid neoplasms based on methylation patterns that, hinged on genomic data, could refine personalized management of thyroid neoplasms.

## Data Availability

All the data used in our study are publicly available at Gene Expression Omnibus (GEO) or GDC Data Portal. All R packages employed in our study are also freely available at CRAN or Bioconductor.

## Notes

### Ethics approval statement

The present study was approved by the institutional ethical committee (Number 47605421400005327).

### CRediT author statement

**V.R.M.:** Conceptualization, methodology, formal analysis, investigation, data curation, writing - original draft, visualization. **M.R.M**.: Methodology, Formal analysis, Data Curation. **A.L.M**.: Investigation, Writing - Review & Editing. **I.M.G**.: Conceptualization, methodology, investigation, writing - review & editing, supervision, project administration. All authors read and approved the final manuscript.

### Prior presentations

These results have not been presented previously.

## Acknowledgments

The results published herein are in whole or part based upon data generated in previous studies and deposited in GEO. The authors thank the patients who participated in the studies. We also thank all personnel involved in these projects for their vast contributions.

## Data Availability

All the data used in our study are publicly available at GEO. All R packages employed in our study are also freely available at CRAN or Bioconductor.

## Supplementary Data

**Supplementary Figure 1:** Flowchart illustrating the search strategy used to select studies eligible for inclusion

**Supplementary Figure 2:** Stability metrics (RCSI and Monte Carlo p value) obtained from spectral consensus clustering using the 10,000 or 20,000 most variable beta-values.

**Supplementary Figure 3:** t-SNE plots employed to inspect cluster separation in the TCGA dataset.

**Supplementary Table 1:** Description of histological subtypes within the discovery dataset.

**Supplementary Table 2:** Association between transduction pathways and differentially methylated positions in PTC-like samples

**Supplementary Table 3:** Association between transduction pathways and differentially methylated positions in follicular-like samples

**Supplementary File 1:** Genomic positions and overlapping genes for the 136 differentially methylated regions in PTC-like samples

**Supplementary File 2:** Genomic positions and overlapping genes for the 57 differentially methylated regions in follicular-like samples

