## Supplementary Data for "DNA methylation patterns define subtypes of differentiated follicular cell-derived thyroid neoplasms: an unsupervised machine learning approach"

### Table of Contents

Supplementary Figure 1

Supplementary Figure 2

Supplementary Figure 3

Supplementary Table 1

Supplementary Table 2

Supplementary Table 3

**Supplementary Figure 1**

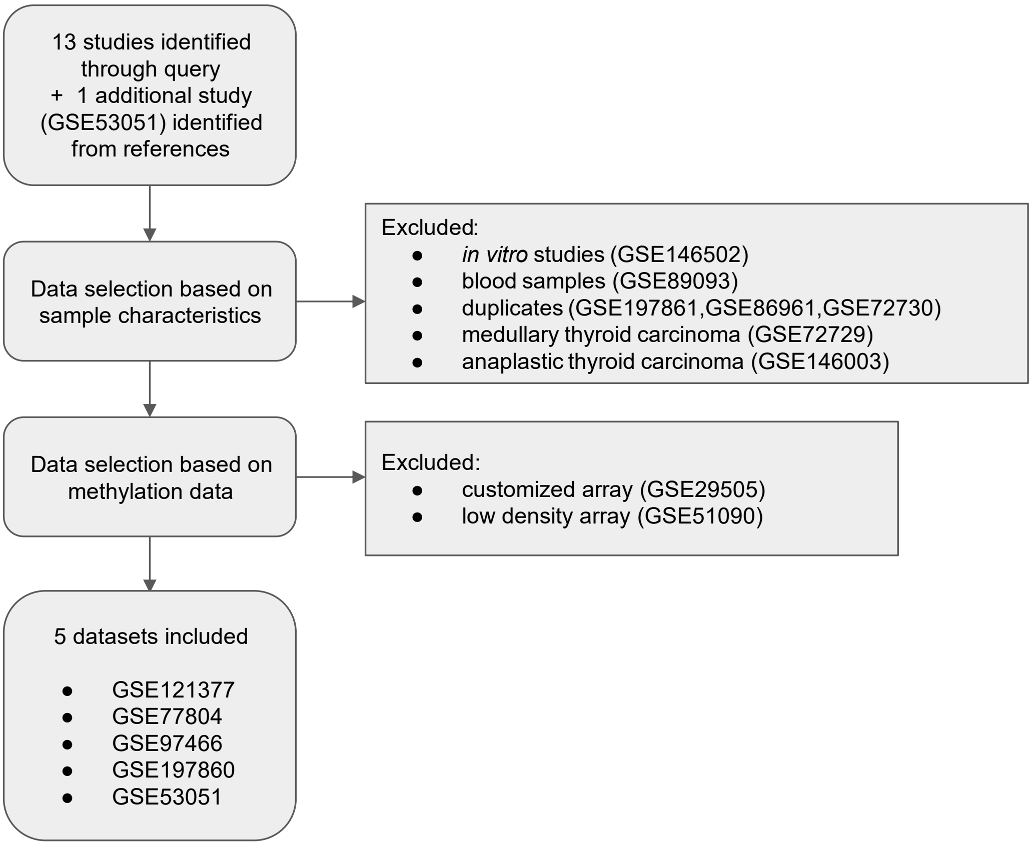


**Supplementary Figure 1:** Flowchart illustrating the search strategy used to select studies eligible for inclusion

**Supplementary Figure 2**

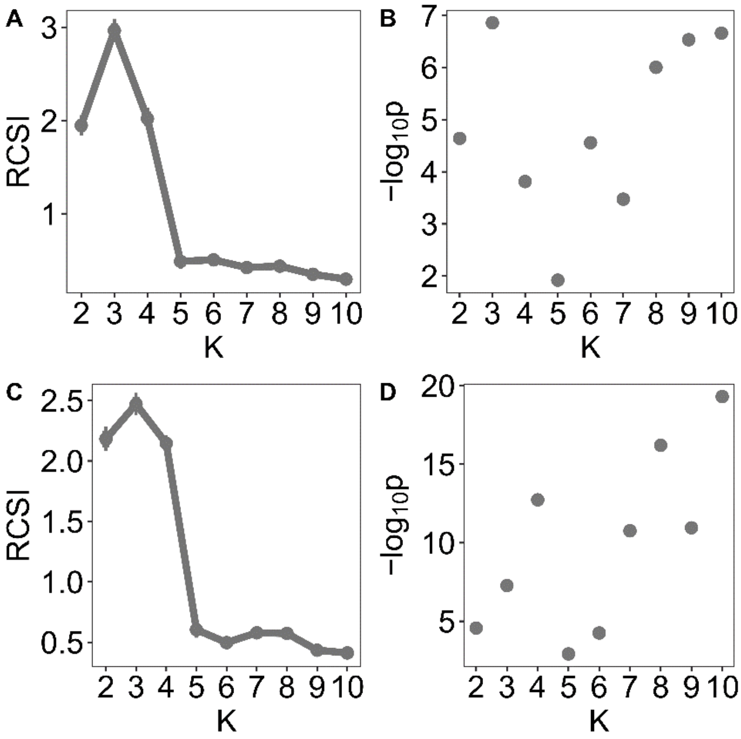


**Supplementary Figure 2:** Stability metrics (RCSI and Monte Carlo p value) obtained from spectral consensus clustering using the 10,000 (A,B) or 20,000 (C,D) most variable beta-values. Abbreviations: RCSI, Relative Cluster Stability Index.

**Supplementary Figure 3**

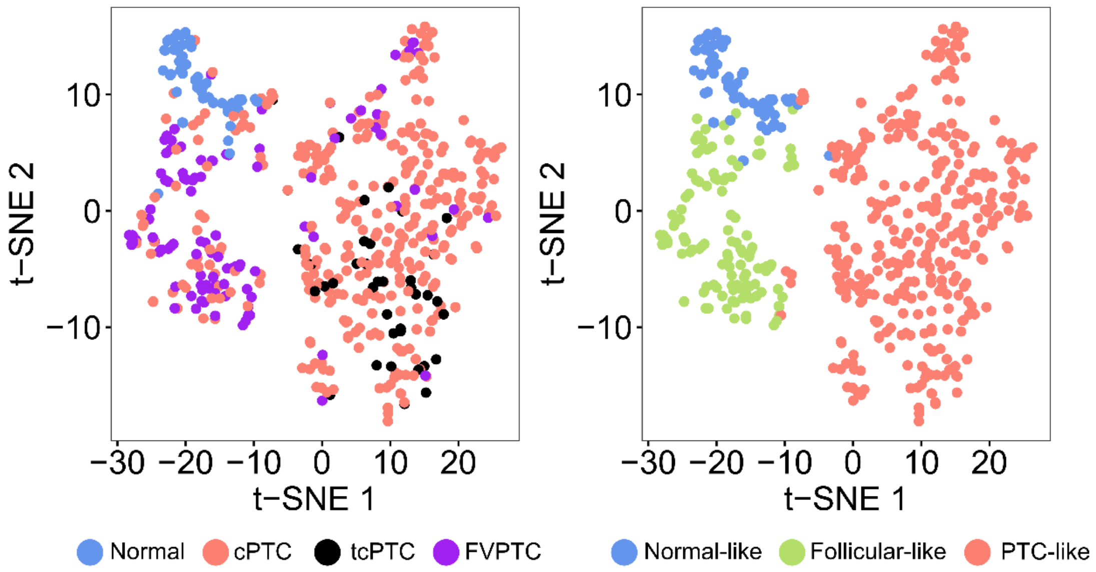


**Supplementary Figure 3:** t-SNE plots were employed to inspect cluster separation in the TCGA dataset. Samples were colored (A) according to histological subtype or (B) according to cluster assignment. Plots were constructed using the 5,000 most variable features from the discovery phase. Abbreviations: cPTC, classic papillary thyroid carcinoma; FVPTC, follicular variant papillary thyroid carcinoma; tcPTC, tall cell papillary thyroid carcinoma; TCGA, The Cancer Genome Atlas; t-SNE, t-distributed stochastic neighbor embedding.

**Supplementary Table 1:** Histological subtypes

| | Number of samples n | (%) |
|---|---|---|
| Normal thyroid | 69 | (27.0) |
| Classic PTC | 70 | (27.3) |
| Tall cell PTC | 8 | (3.1) |
| Follicular variant PTC | 21 | (8.2) |
| Follicular thyroid carcinoma | 22 | (8.6) |
| Follicular adenoma | 16 | (6.3) |
| NIFTP | 6 | (2.3) |
| Oncocytic Carcinoma | 16 | (6.3) |
| Oncocytic Adenoma | 16 | (6.3) |
| Goiter | 12 | (4.7) |
| **Total** | **256** | |

**Abbreviations:** PTC, papillary thyroid carcinoma; NIFTP, noninvasive follicular thyroid neoplasm with papillary-like nuclear features.

**Supplementary Table 2:** Association between transduction pathways and differentially methylated positions in PTC-like samples

| Pathway ID | Pathway Name | p-value | Adjusted p-value |
|---|---|---|---|
| R-HSA-2219528 | PI3K/AKT Signaling in Cancer | <0.0001 | 0.0002 |
| R-HSA-194138 | Signaling by VEGF | <0.0001 | 0.0002 |
| R-HSA-5683057 | MAPK family signaling cascades | <0.0001 | 0.0002 |
| R-HSA-2219530 | Constitutive Signaling by Aberrant PI3K in Cancer | 0.0001 | 0.0004 |
| R-HSA-5684996 | MAPK1/MAPK3 signaling | 0.0003 | 0.0018 |
| R-HSA-6806834 | Signaling by MET | 0.0005 | 0.0029 |
| R-HSA-166520 | Signaling by NTRKs | 0.0022 | 0.0092 |
| R-HSA-9006925 | Intracellular signaling by second messengers | 0.0023 | 0.0092 |
| R-HSA-1257604 | PIP3 activates AKT signaling | 0.0102 | 0.0364 |
| R-HSA-193704 | p75 NTR receptor-mediated signalling | 0.0463 | 0.1481 |
| R-HSA-1980143 | Signaling by NOTCH1 | 0.0813 | 0.2364 |
| R-HSA-73887 | Death Receptor Signalling | 0.1040 | 0.2774 |
| R-HSA-74752 | Signaling by Insulin receptor | 0.1241 | 0.3023 |
| R-HSA-157118 | Signaling by NOTCH | 0.1342 | 0.3023 |
| R-HSA-195721 | Signaling by WNT | 0.1417 | 0.3023 |
| R-HSA-1226099 | Signaling by FGFR in disease | 0.1924 | 0.3848 |
| R-HSA-6802957 | Oncogenic MAPK signaling | 0.2346 | 0.4416 |
| R-HSA-9006931 | Signaling by Nuclear Receptors | 0.2863 | 0.4974 |
| R-HSA-8939211 | ESR-mediated signaling | 0.3008 | 0.4974 |
| R-HSA-190236 | Signaling by FGFR | 0.3109 | 0.4974 |
| R-HSA-3858494 | Beta-catenin independent WNT signaling | 0.5425 | 0.7968 |
| R-HSA-6802952 | Signaling by BRAF and RAF1 fusions | 0.5478 | 0.7968 |
| R-HSA-201681 | TCF dependent signaling in response to WNT | 0.7212 | 0.9875 |
| R-HSA-5687128 | MAPK6/MAPK4 signaling | 0.8171 | 0.9875 |
| R-HSA-5358351 | Signaling by Hedgehog | 0.8761 | 0.9875 |
| R-HSA-5632684 | Hedgehog 'on' state | 0.8946 | 0.9875 |
| R-HSA-9006936 | Signaling by TGFB family members | 0.9381 | 0.9875 |
| R-HSA-195253 | Degradation of beta-catenin by the destruction complex | 0.9400 | 0.9875 |
| R-HSA-9013694 | Signaling by NOTCH4 | 0.9400 | 0.9875 |
| R-HSA-5610787 | Hedgehog 'off' state | 0.9469 | 0.9875 |
| R-HSA-170834 | Signaling by TGF-beta Receptor Complex | 0.9566 | 0.9875 |
| R-HSA-5358346 | Hedgehog ligand biogenesis | 0.9992 | 0.9992 |

**Supplementary Table 3:** Association between transduction pathways and differentially methylated positions in follicular-like samples

| Pathway ID | Pathway Name | p-value | Adjusted p-value |
|---|---|---|---|
| R-HSA-170834 | Signaling by TGF-beta Receptor Complex | <0.0001 | <0.0001 |
| R-HSA-193704 | p75 NTR receptor-mediated signalling | <0.0001 | <0.0001 |
| R-HSA-194138 | Signaling by VEGF | <0.0001 | <0.0001 |
| R-HSA-1980143 | Signaling by NOTCH1 | <0.0001 | <0.0001 |
| R-HSA-6802952 | Signaling by BRAF and RAF1 fusions | <0.0001 | <0.0001 |
| R-HSA-6802957 | Oncogenic MAPK signaling | <0.0001 | <0.0001 |
| R-HSA-73887 | Death Receptor Signalling | <0.0001 | <0.0001 |
| R-HSA-9006936 | Signaling by TGFB family members | <0.0001 | <0.0001 |
| R-HSA-5683057 | MAPK family signaling cascades | 0.0136 | 0.0459 |
| R-HSA-166520 | Signaling by NTRKs | 0.0143 | 0.0459 |
| R-HSA-5684996 | MAPK1/MAPK3 signaling | 0.0291 | 0.0848 |
| R-HSA-157118 | Signaling by NOTCH | 0.0324 | 0.0863 |
| R-HSA-9006925 | Intracellular signaling by second messengers | 0.0473 | 0.1165 |
| R-HSA-5358351 | Signaling by Hedgehog | 0.0571 | 0.1305 |
| R-HSA-6806834 | Signaling by MET | 0.0802 | 0.1642 |
| R-HSA-74752 | Signaling by Insulin receptor | 0.0855 | 0.1642 |
| R-HSA-195721 | Signaling by WNT | 0.0872 | 0.1642 |
| R-HSA-201681 | TCF dependent signaling in response to WNT | 0.1032 | 0.1740 |
| R-HSA-1257604 | PIP3 activates AKT signaling | 0.1033 | 0.1740 |
| R-HSA-5610787 | Hedgehog 'off' state | 0.1417 | 0.2268 |
| R-HSA-2219528 | PI3K/AKT Signaling in Cancer | 0.1647 | 0.2509 |
| R-HSA-5632684 | Hedgehog 'on' state | 0.2607 | 0.3442 |
| R-HSA-5687128 | MAPK6/MAPK4 signaling | 0.2669 | 0.3442 |
| R-HSA-8939211 | ESR-mediated signaling | 0.2748 | 0.3442 |
| R-HSA-195253 | Degradation of beta-catenin by the destruction complex | 0.2796 | 0.3442 |
| R-HSA-9013694 | Signaling by NOTCH4 | 0.2796 | 0.3442 |
| R-HSA-2219530 | Constitutive Signaling by Aberrant PI3K in Cancer | 0.2997 | 0.3552 |
| R-HSA-3858494 | Beta-catenin independent WNT signaling | 0.3462 | 0.3956 |
| R-HSA-5358346 | Hedgehog ligand biogenesis | 0.4007 | 0.4422 |
| R-HSA-9006931 | Signaling by Nuclear Receptors | 0.4288 | 0.4574 |
| R-HSA-190236 | Signaling by FGFR | 0.4982 | 0.5143 |
| R-HSA-1226099 | Signaling by FGFR in disease | 0.7025 | 0.7025 |
